# Specialty Organization Finances and Membership: A Closer Look

**DOI:** 10.1101/2023.09.16.23295655

**Authors:** Ali Syed, Ehsan Rahimy, Jayanth Sridhar

## Abstract

**Background:** United States students and physicians face massive financial hurdles including student loan repayments, board certification fees, and other expenses. This project investigates an infrequently discussed topic, the costs to join and the finances of national specialty organizations.

**Purpose:** We hope our findings encourage resident physicians and newly minted attendings to take interest in the handling of the fees they may pay to join a national organization.

**Methods:** Publicly available IRS 990 filings and specialty organization websites were examined to compile data regarding membership fees, organization revenue, expenses, and top earner salaries for 29 total specialty organizations. Descriptive and inferential statistics including Pearson correlation coefficients were used to investigate the above data.

**Results:** The average annual revenue and net assets for the organizations studied were $50,762,279 and $66,444,291, respectively. The average annual membership fee was $542. The largest ratios of membership fee to physician salary were in pediatrics (0.31%) and ophthalmology (0.26%). The average salary of the top three earners at specialty organizations was $429,758.

**Conclusions:** There is wide variability in the costs of joining medical specialty organizations in the United States that does not correlate with the average earning potential of individual specialties. As a whole, medical specialty organizations should consider greater financial transparency.

## Text

Early in their career, United States physicians face a broad number of financial pressures. For one data point, the median student debt coming out of United States medical schools in 2019 was $200,000.^1^ After investing significant sums of money in medical school, board exams, and state licensure, many physicians finishing training come to realize that American medicine is a pay-to-play game. Even after being well-established as full-fledged attendings, U.S. physicians face even more financial obstacles including licensure renewal, maintenance of board certification fees, and the costs of joining their respective specialty organizations. In 2017, Drolet and Tandon investigated some of these financial obstacles.^2^ They found that the mean fee for an initial written examination for board certification was $1863 across 24 specialties. In addition, 14 of these boards required an oral examination at a mean cost of $1695. And even after attaining initial certification, a mean annual fee of $262 was required for maintenance of certification. This study clearly delineated the initial and maintenance costs of certification. However, there are still many topics relating to the costs of being a physician that have not yet been well investigated. To our knowledge, there has not been a study that consolidates financial data and costs of membership from medical specialty organizations. In this viewpoint, we describe and provide commentary related to our findings including membership fees, organization revenue and expenses, and the salaries of top earners at medical specialty organizations.

## Methods

To objectively assess, we extracted specialty organization revenue, expenses, and net assets data from nonprofitlight.com which consolidates data from IRS 990 filings of 501(c)(3) organizations. We extracted this publicly available data from 29 different national medical societies, corresponding to 29 different medical specialties. Although most specialties have multiple organizations, we chose to represent each specialty using the financial data from one of their prominent societies. Membership data, including annual membership fees, were taken from specialty organization websites. Membership fees vary according to the degree of training. Only membership fees for attending-level physicians were used for analysis.

## Results

The average annual revenue and net assets for these 29 organizations were $50,762,279 and $66,444,291, respectively (Table 1). The average annual membership fee for attending-level physicians was $580. There is a moderate correlation (r = 0.52) between the average physician salary in each individual specialty and the annual membership fee of the respective specialty organization. Dividing the annual membership fee by the average physician salary within the specialty, taken from Medscape’s 2021 Physician Compensation Report, gives insight into which organizations are the most and least expensive to join relative to the annual salary of their members.^3^ The largest ratios are present in pediatrics (0.31%) and ophthalmology (0.26%), while the lowest ratios are seen in endocrinology (0.08%) and obstetrics and gynecology (0.08%). The number of board-certified members was ascertained from eleven of these organizations, and there were strong correlations between the number of board-certified members and specialty organization revenue (r = 0.858, p < 0.001) and net assets (r = 0.673, p = 0.02).

We also extracted the salaries, roles, and educational backgrounds of the top three earners from each of these organizations. Across all organizations, the top earners were either c-suite executives or organization vice presidents. We gave a single-degree designation to each of these top earners. Priority was given to more advanced degrees if an individual held multiple degrees (e.g. an individual who has an MD, MBA, and BS was classified as an MD). There was a strong correlation (r = 0.839, p < 0.001) between organization revenue and the average salaries of the top three earners within the organization. The average salary for the top earners across the 29 organizations was $429,758. These leaders come from a wide variety of backgrounds with the plurality of them holding either an MD or DO degree (30%), but the mix includes 10% JDs, 8% PhDs (or other doctoral), 17% MBAs, and 29% BS/BA or other. The salaries of these leaders did differ depending on the degree type. A t-test assuming unequal variance between MD/DOs and MBAs found a significant difference in salary ($537,367 vs $350,127 p = 0.0053). A t-test assuming unequal variance between MD/DOs and BS/BAs found a significant difference in salary ($537,367 vs $329,489 p = 0.0015). ANOVA showed no difference between the average salary of top earners with different doctorate-level degrees (i.e., MD/DOs vs JD vs Ph.D. or other doctorates).

## Discussion

Our findings consolidate and provide a general overview of specialty organization finances including organization revenue, net assets, and membership fees. Although limited data may be available through organization annual reports, these reports generally do not include more detailed information such as salaries for organization leaders. The 2022 Medscape Physician Compensation Report found that physician salaries overall average $339,000.^4^ The average compensation of top earners at specialty organizations is 27% greater. Although the role of a physician is significantly different than that of a specialty organization leader, we believe it is important for both parties to be aware of each other’s compensation.

We also found that annual membership fees are variable, ranging from $205 to $1,076. Depending on the average salary within a specialty, these fees may be more or less affordable for physician members. We found that membership was most expensive relative to salary in ophthalmology and pediatrics and least expensive in endocrinology and obstetrics and gynecology. However, this analysis does not take into consideration the bundle of goods that each organization provides to its members. Organizations may use the revenue generated from membership fees to provide a variety of benefits to their members including journal subscriptions, loan refinancing plans, networking events, and support on advocacy issues.

Recently, the American Academy of Ophthalmology (AAO) and the American Society of Cataract and Refractive Surgery (ASCRS) mounted a public relations campaign against Aetna for their use of blanket prior authorizations for cataract surgery. This requirement was ultimately dropped on July 1^st^, 2022, for all members except Florida and Georgia Medicare Advantage members.^5^ The unique benefits each organization provides suggest that it is difficult to objectively assess how valuable an organization is to its members.

In the process of collecting data, we also saw examples of organizations considering the financial situation of their physician members. Some organizations, such as the American College of Emergency Physicians (ACEP), have unique due structures for newly graduated residents. However, only 10 of the 29 organizations (34%) offer these discounted due structures. Discounted due structures for newly graduated residents provide some financial relief at a time when work-related expenses such as board certification fees are high. We support discounted due structures for newly minted attendings and believe they may encourage early involvement in specialty organizations.

As we continue to scrutinize the financial aspects of medicine, we should find value in investigating the spaces that directly impact physicians. As attendings get inundated with opportunities to spend their hard-earned money, it is important that they are aware to some degree of the finances behind these organizations. We encourage all specialty organization members to look at their specialty organization’s annual report, if available, to gain a rough understanding of where money is coming from and where it is going. We also encourage all organizations to provide more granular financial information for their members. Not all organizations consolidate their financials in an annual report. Broader adoption of this practice would be a step in the right direction. We hope this study spurs further analyses relating to specialty organizations, including their capital allocation.

## Data Availability

All data produced in the present study are available upon reasonable request to the authors

## Financial and Membership Data of Specialty Organizations

**Table S1:**
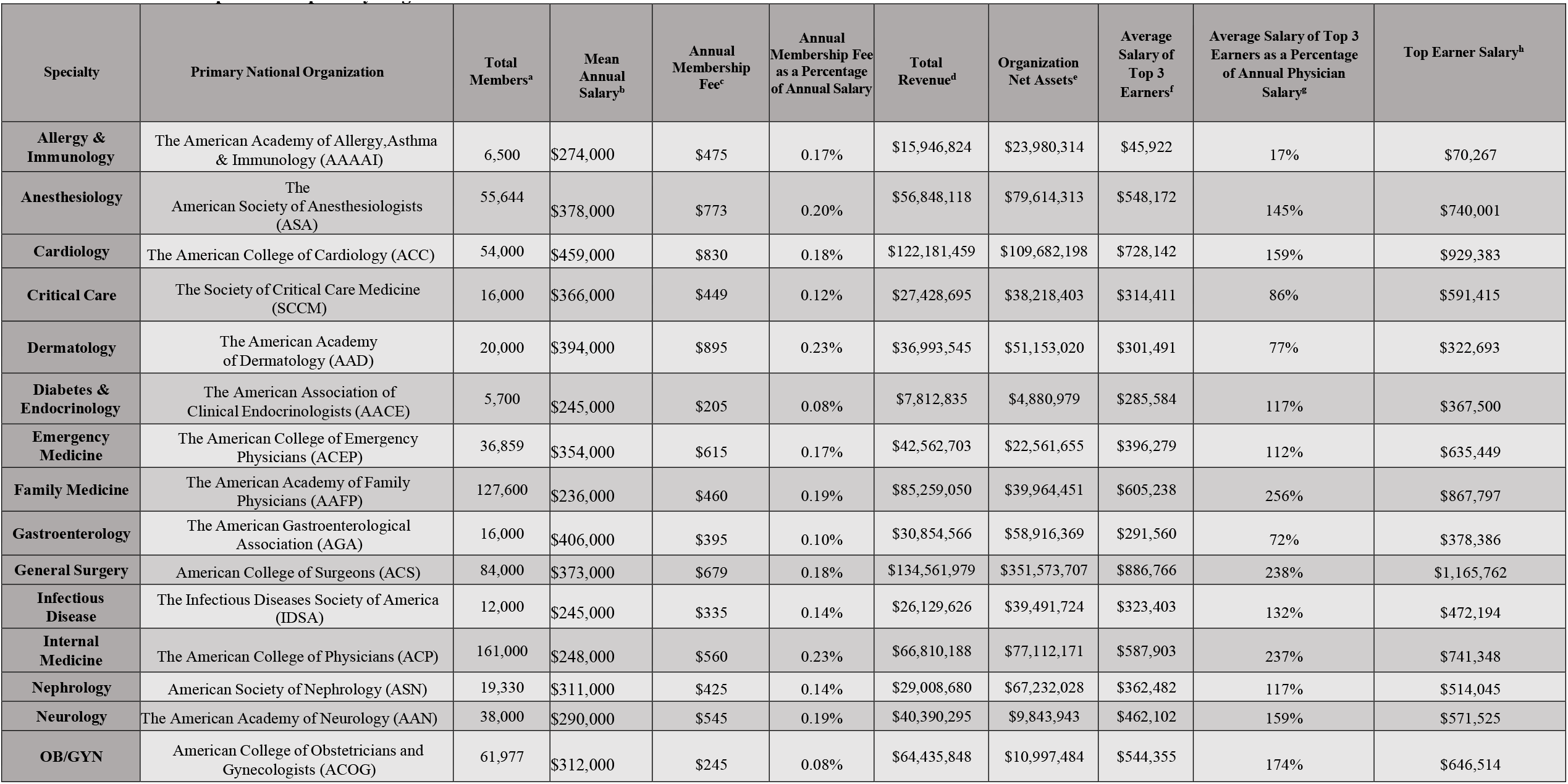
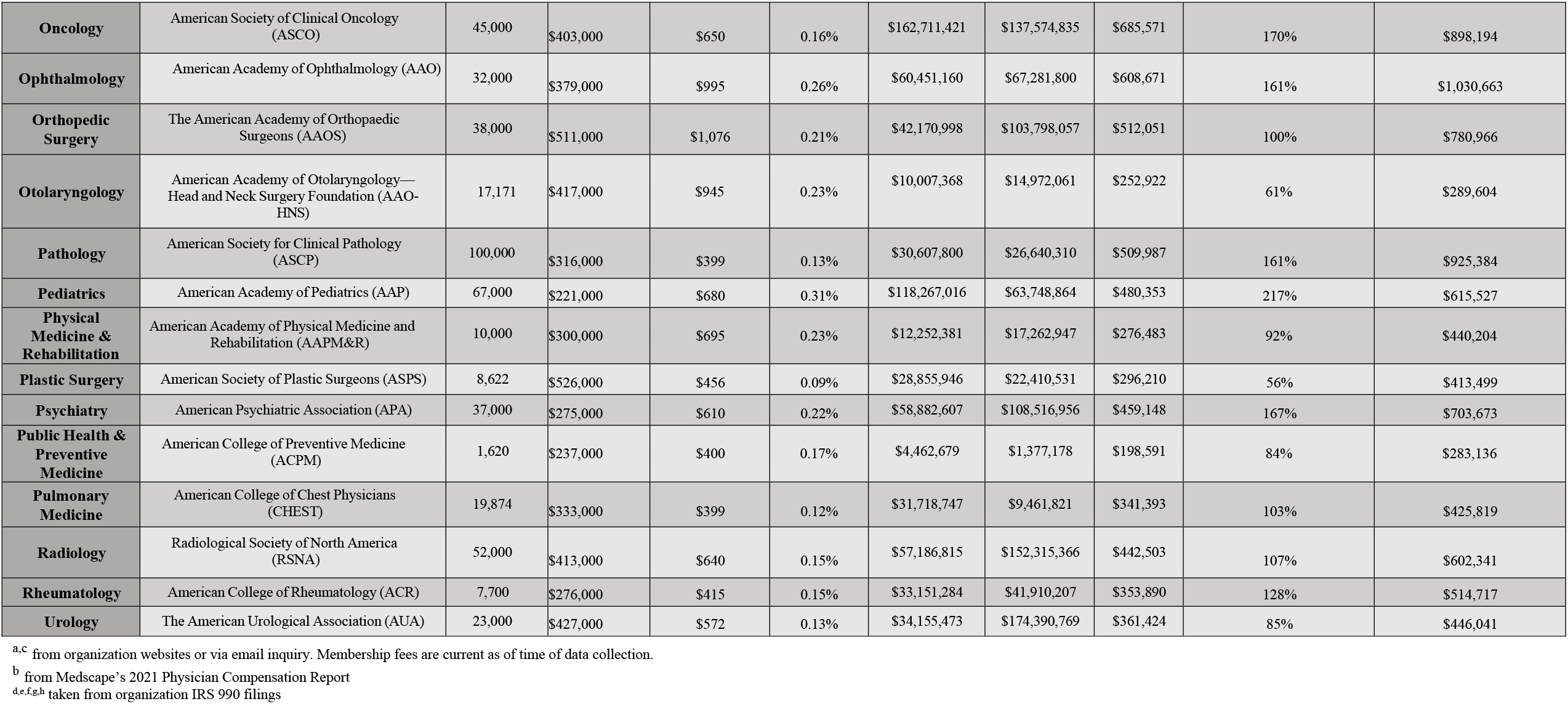
The table above displays the twenty-nine medical specialties of interest, the average salary of physicians in the specialty, the name of the leading specialty organization, membership and financial data of the specialty organization, and the annual membership fee for attending-level physicians to join the specialty organization.

| Specialty | Primary National Organization | Total Members<sup>a</sup> | Mean Annual Salary<sup>b</sup> | Annual Membership Fee<sup>c</sup> | Annual Membership Fee as a Percentage of Annual Salary | Total Revenue<sup>d</sup> | Organization Net Assets<sup>e</sup> | Average Salary of Top 3 Earners<sup>f</sup> | Average Salary of Top 3 Earners as a Percentage of Annual Physician Salary<sup>g</sup> | Top Earner Salary<sup>h</sup> |
| --- | --- | --- | --- | --- | --- | --- | --- | --- | --- | --- |
| Allergy & Immunology | The American Academy of Allergy,Asthma & Immunology (AAAAI) | 6,500 | \$274,000 | \$475 | 0.17% | \$15,946,824 | \$23,980,314 | \$45,922 | 17% | \$70,267 |
| Anesthesiology | The American Society of Anesthesiologists (ASA) | 55,644 | \$378,000 | \$773 | 0.20% | \$56,848,118 | \$79,614,313 | \$548,172 | 145% | \$740,001 |
| Cardiology | The American College of Cardiology (ACC) | 54,000 | \$459,000 | \$830 | 0.18% | \$122,181,459 | \$109,682,198 | \$728,142 | 159% | \$929,383 |
| Critical Care | The Society of Critical Care Medicine (SCCM) | 16,000 | \$366,000 | \$449 | 0.12% | \$27,428,695 | \$38,218,403 | \$314,411 | 86% | \$591,415 |
| Dermatology | The American Academy of Dermatology (AAD) | 20,000 | \$394,000 | \$895 | 0.23% | \$36,993,545 | \$51,153,020 | \$301,491 | 77% | \$322,693 |
| Diabetes & Endocrinology | The American Association of Clinical Endocrinologists (AACE) | 5,700 | \$245,000 | \$205 | 0.08% | \$7,812,835 | \$4,880,979 | \$285,584 | 117% | \$367,500 |
| Emergency Medicine | The American College of Emergency Physicians (ACEP) | 36,859 | \$354,000 | \$615 | 0.17% | \$42,562,703 | \$22,561,655 | \$396,279 | 112% | \$635,449 |
| Family Medicine | The American Academy of Family Physicians (AAFP) | 127,600 | \$236,000 | \$460 | 0.19% | \$85,259,050 | \$39,964,451 | \$605,238 | 256% | \$867,797 |
| Gastroenterology | The American Gastroenterological Association (AGA) | 16,000 | \$406,000 | \$395 | 0.10% | \$30,854,566 | \$58,916,369 | \$291,560 | 72% | \$378,386 |
| General Surgery | American College of Surgeons (ACS) | 84,000 | \$373,000 | \$679 | 0.18% | \$134,561,979 | \$351,573,707 | \$886,766 | 238% | \$1,165,762 |
| Infectious Disease | The Infectious Diseases Society of America (IDSA) | 12,000 | \$245,000 | \$335 | 0.14% | \$26,129,626 | \$39,491,724 | \$323,403 | 132% | \$472,194 |
| Internal Medicine | The American College of Physicians (ACP) | 161,000 | \$248,000 | \$560 | 0.23% | \$66,810,188 | \$77,112,171 | \$587,903 | 237% | \$741,348 |
| Nephrology | American Society of Nephrology (ASN) | 19,330 | \$311,000 | \$425 | 0.14% | \$29,008,680 | \$67,232,028 | \$362,482 | 117% | \$514,045 |
| Neurology | The American Academy of Neurology (AAN) | 38,000 | \$290,000 | \$545 | 0.19% | \$40,390,295 | \$9,843,943 | \$462,102 | 159% | \$571,525 |
| OB/GYN | American College of Obstetricians and Gynecologists (ACOG) | 61,977 | \$312,000 | \$245 | 0.08% | \$64,435,848 | \$10,997,484 | \$544,355 | 174% | \$646,514 |
| **Oncology** | American Society of Clinical Oncology (ASCO) | 45,000 | \$403,000 | \$650 | 0.16% | \$162,711,421 | \$137,574,835 | \$685,571 | 170% | \$898,194 |
| **Ophthalmology** | American Academy of Ophthalmology (AAO) | 32,000 | \$379,000 | \$995 | 0.26% | \$60,451,160 | \$67,281,800 | \$608,671 | 161% | \$1,030,663 |
| **Orthopedic Surgery** | The American Academy of Orthopaedic Surgeons (AAOS) | 38,000 | \$511,000 | \$1,076 | 0.21% | \$42,170,998 | \$103,798,057 | \$512,051 | 100% | \$780,966 |
| **Otolaryngology** | American Academy of Otolaryngology—Head and Neck Surgery Foundation (AAO-HNS) | 17,171 | \$417,000 | \$945 | 0.23% | \$10,007,368 | \$14,972,061 | \$252,922 | 61% | \$289,604 |
| **Pathology** | American Society for Clinical Pathology (ASCP) | 100,000 | \$316,000 | \$399 | 0.13% | \$30,607,800 | \$26,640,310 | \$509,987 | 161% | \$925,384 |
| **Pediatrics** | American Academy of Pediatrics (AAP) | 67,000 | \$221,000 | \$680 | 0.31% | \$118,267,016 | \$63,748,864 | \$480,353 | 217% | \$615,527 |
| **Physical Medicine & Rehabilitation** | American Academy of Physical Medicine and Rehabilitation (AAPM&R) | 10,000 | \$300,000 | \$695 | 0.23% | \$12,252,381 | \$17,262,947 | \$276,483 | 92% | \$440,204 |
| **Plastic Surgery** | American Society of Plastic Surgeons (ASPS) | 8,622 | \$526,000 | \$456 | 0.09% | \$28,855,946 | \$22,410,531 | \$296,210 | 56% | \$413,499 |
| **Psychiatry** | American Psychiatric Association (APA) | 37,000 | \$275,000 | \$610 | 0.22% | \$58,882,607 | \$108,516,956 | \$459,148 | 167% | \$703,673 |
| **Public Health & Preventive Medicine** | American College of Preventive Medicine (ACPM) | 1,620 | \$237,000 | \$400 | 0.17% | \$4,462,679 | \$1,377,178 | \$198,591 | 84% | \$283,136 |
| **Pulmonary Medicine** | American College of Chest Physicians (CHEST) | 19,874 | \$333,000 | \$399 | 0.12% | \$31,718,747 | \$9,461,821 | \$341,393 | 103% | \$425,819 |
| **Radiology** | Radiological Society of North America (RSNA) | 52,000 | \$413,000 | \$640 | 0.15% | \$57,186,815 | \$152,315,366 | \$442,503 | 107% | \$602,341 |
| **Rheumatology** | American College of Rheumatology (ACR) | 7,700 | \$276,000 | \$415 | 0.15% | \$33,151,284 | \$41,910,207 | \$353,890 | 128% | \$514,717 |
| **Urology** | The American Urological Association (AUA) | 23,000 | \$427,000 | \$572 | 0.13% | \$34,155,473 | \$174,390,769 | \$361,424 | 85% | \$446,041 |

<sup>a,c</sup> from organization websites or via email inquiry. Membership fees are current as of time of data collection.
<sup>b</sup> from Medscape’s 2021 Physician Compensation Report
<sup>d,e,f,g,h</sup> taken from organization IRS 990 filings

## Notes

### Competing Interest Statement

The authors have declared no competing interest.

### Funding Statement

This study did not receive any funding

